## Supplementary material for "The effect of previous SARS-CoV-2 infection and COVID-19 vaccination on SARS-CoV-2 Omicron infection and relation with serological response – a prospective cohort study": Figure S

To manuscript

*Table S1. Vaccines received by study participants, per dose. Participants who did not receive the dose were excluded from the percentage calculation.*

| | | **N (%)** |
|---|---|---|
| **Dose 1** | AstraZeneca (Vaxzevria) | 14585 (34.5) |
| | BioNTech/ Pfizer (Comirnaty) | 17036 (40.3) |
| | Janssen | 4729 (11.2) |
| | Moderna (Spikevax) | 5864 (13.9) |
| | Unknown | 31 ( 0.1) |
| **Dose 2** | AstraZeneca (Vaxzevria) | 13505 (32.3) |
| | BioNTech/ Pfizer (Comirnaty) | 19191 (45.9) |
| | Janssen | 115 ( 0.3) |
| | Moderna (Spikevax) | 7971 (19.0) |
| | Novavax | 2 ( 0.0) |
| | Unknown | 1072 ( 2.6) |
| **Dose 3** | AstraZeneca (Vaxzevria) | 343 ( 1.0) |
| | BioNTech/ Pfizer (Comirnaty) | 13822 (38.5) |
| | Janssen | 18 ( 0.1) |
| | Moderna (Spikevax) | 21408 (59.7) |
| | Novavax | 2 ( 0.0) |
| | Unknown | 264 ( 0.7) |
| **Dose 4** | AstraZeneca (Vaxzevria) | 99 ( 0.8) |
| | BioNTech/ Pfizer (Comirnaty) | 3239 (27.3) |
| | Janssen | 4 ( 0.0) |
| | Moderna (Spikevax) | 8159 (68.7) |
| | Novavax | 5 ( 0.0) |
| | Unknown | 374 ( 3.1) |

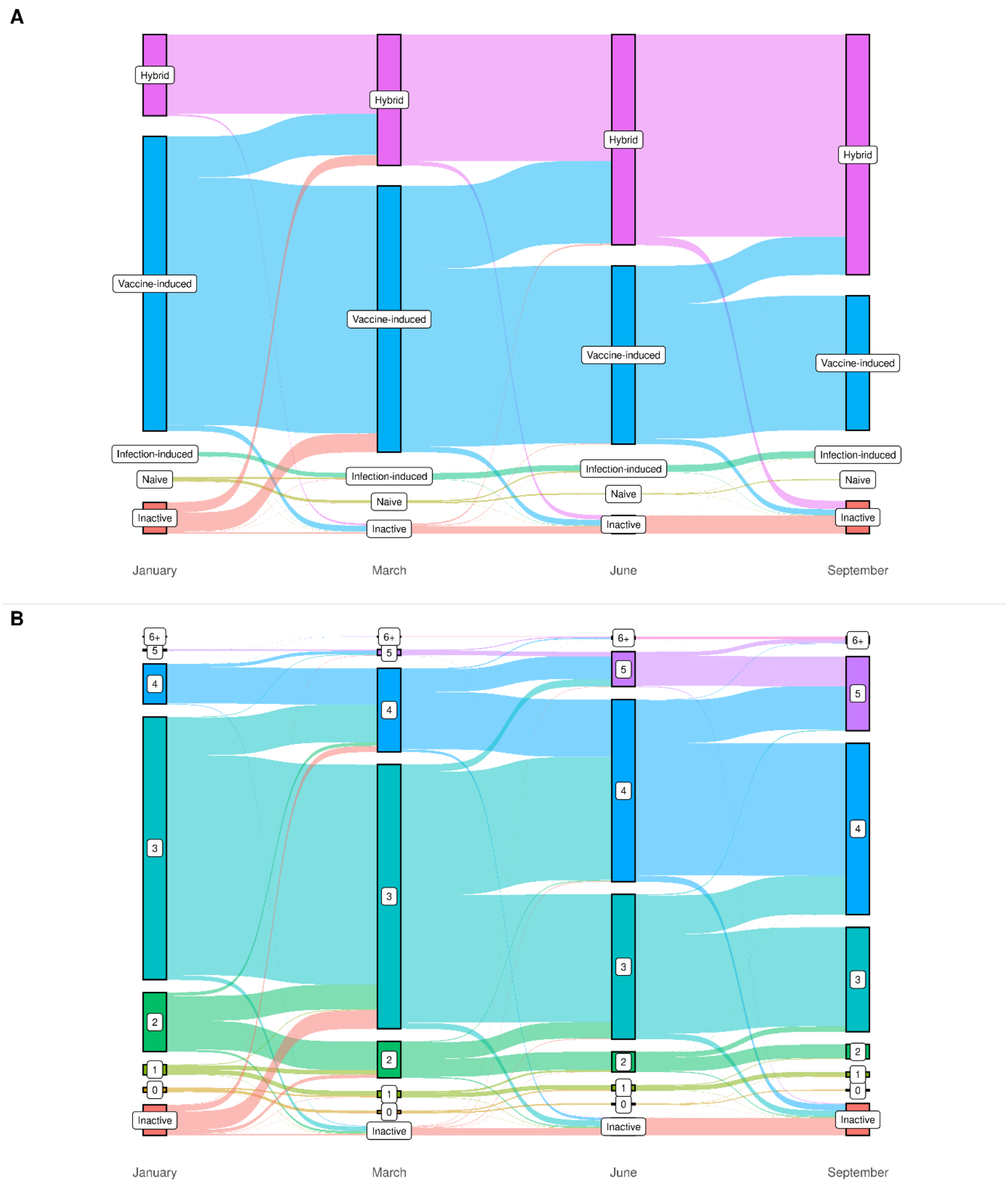


*Figure S1. Sankey diagrams showing the changes in the VASCO study population during the study period (January-September 2022), in terms of type of immunity (A) and number of prior immunizing events (B).*

*Table S2. Number of person-days and SARS-CoV-2 infections during the study period, adjusted hazard ratios (aHR) and 95% confidence intervals (CI) for SARS-CoV-2 infection, number of serology samples available, adjusted geometric mean S-antibody concentration (GMC) and adjusted GMC ratio, by type of immunity and number of exposures. Four to 10 weeks after Vaccine-induced served as the reference group*. The group with 4 exposures includes only participants aged 60 and older, because younger people were not eligible for 4 vaccinations.

| Type of immunity | Weeks since last immunizing event | Number of immunizing events | Person-days | Infections | aHR (95% CI) | Number of serology samples | Geometric mean anti-S (95%CI) | GMC ratio (95%CI) |
|---|---|---|---|---|---|---|---|---|
| **Vaccine-induced** | [4,10) | 2 | 108554 | 708 | [ref] | 237 | 8915 (7973-9968) | [ref] |
| **Vaccine-induced** | [10,20) | 2 | 127890 | 810 | 1.14 (1.01-1.28) | 181 | 4709 (4180-5306) | 0.52 (0.42-0.65) |
| **Vaccine-induced** | [20,30) | 2 | 107641 | 497 | 1.52 (1.31-1.76) | 776 | 3125 (2912-3353) | 0.33 (0.28-0.39) |
| **Vaccine-induced** | [30,40] | 2 | 76420 | 371 | 1.38 (1.19-1.59) | 132 | 1722 (1279-2318) | 0.19 (0.15-0.25) |
| **Hybrid** | [4,10) | 2 | 12832 | 12 | 0.16 (0.09-0.28) | 28 | 15028 (9786-23077) | 1.76 (1.13-2.75) |
| **Hybrid** | [10,20) | 2 | 20102 | 54 | 0.52 (0.39-0.69) | 70 | 9487 (7583-11869) | 1.07 (0.79-1.45) |
| **Hybrid** | [20,30) | 2 | 11964 | 15 | 0.40 (0.24-0.67) | 44 | 6132 (4407-8531) | 0.70 (0.48-1.0) |
| **Hybrid** | [30,40] | 2 | 6804 | 19 | 0.72 (0.46-1.15) | 18 | 6003 (3884-9277) | 0.69 (0.4-1.2) |
| **Infection-induced** | [4,10) | 2 | 11542 | 3 | 0.06 (0.02-0.17) | 44 | 1650 (868-3136) | 0.2 (0.14-0.28) |
| **Infection-induced** | [10,20) | 2 | 16173 | 8 | 0.17 (0.08-0.35) | 79 | 1205 (787-1846) | 0.14 (0.1-0.19) |
| **Infection-induced** | [20,30) | 2 | 10753 | 3 | 0.12 (0.04-0.36) | 9 | 3234 (1628-6423) | 0.37 (0.17-0.78) |
| **Infection-induced** | [30,40] | 2 | 1797 | 5 | 1.48 (0.61-3.64) | 2 | 1134 (43-29809) | 0.15 (0.03-0.80) |
| **Vaccine-induced** | [4,10) | 3 | 1021822 | 3738 | [ref] | 2029 | 17361 (16736-18009) | [ref] |
| **Vaccine-induced** | [10,20) | 3 | 1177085 | 5898 | 1.29 (1.22-1.37) | 1056 | 8770 (8190-9391) | 0.52 (0.48-0.56) |
| **Vaccine-induced** | [20,30) | 3 | 711013 | 2033 | 1.89 (1.69-2.11) | 5667 | 8711 (8477-8952) | 0.48 (0.46-0.51) |
| **Vaccine-induced** | [30,40] | 3 | 321224 | 625 | 2.31 (1.99-2.68) | 449 | 6476 (5806-7225) | 0.35 (0.31-0.38) |
| **Hybrid** | [4,10) | 3 | 210792 | 255 | 0.29 (0.26-0.33) | 719 | 23285 (21824-24844) | 1.51 (1.38-1.64) |
| **Hybrid** | [10,20) | 3 | 296881 | 517 | 0.54 (0.48-0.59) | 1265 | 19444 (18468-20472) | 1.32 (1.23-1.42) |
| **Hybrid** | [20,30) | 3 | 170143 | 217 | 0.69 (0.59-0.81) | 729 | 11030 (10257-11860) | 0.72 (0.67-0.79) |
| **Hybrid** | [30,40] | 3 | 53892 | 89 | 1 (0.80-1.25) | 98 | 8750 (6950-11017) | 0.55 (0.45-0.67) |
| **Vaccine-induced** | [4,10) | 4 | 258106 | 384 | [ref] | 1822 | 21147 (20357-21968) | [ref] |
| **Vaccine-induced** | [10,20) | 4 | 323388 | 778 | 1.31 (1.13-1.51) | 1610 | 14685 (14051-15348) | 0.64 (0.61-0.69) |

| | | | | | | | | |
|---|---|---|---|---|---|---|---|---|
| **Vaccine-induced** | [20,30) | 4 | 53415 | 53 | 1.27 (0.93-1.74) | 71 | 11686 (8567-15942) | 0.55 (0.45-0.68) |
| **Vaccine-induced** | [30,40] | 4 | 4090 | 5 | 1.13 (0.46-2.75) | 5 | 5225 (2487-10975) | 0.26 (0.12-0.58) |
| **Hybrid** | [4,10) | 4 | 391503 | 195 | 0.15 (0.12-0.18) | 998 | 35690 (33830-37653) | 1.69 (1.58-1.82) |
| **Hybrid** | [10,20) | 4 | 456237 | 392 | 0.32 (0.27-0.37) | 2744 | 32810 (31680-33980) | 1.52 (1.44-1.61) |
| **Hybrid** | [20,30) | 4 | 170792 | 107 | 0.43 (0.34-0.54) | 805 | 19466 (18159-20868) | 0.9 (0.84-0.97) |
| **Hybrid** | [30,40] | 4 | 36981 | 28 | 0.7 (0.47-1.05) | 38 | 16771 (12600-22323) | 0.76 (0.57-1.02) |

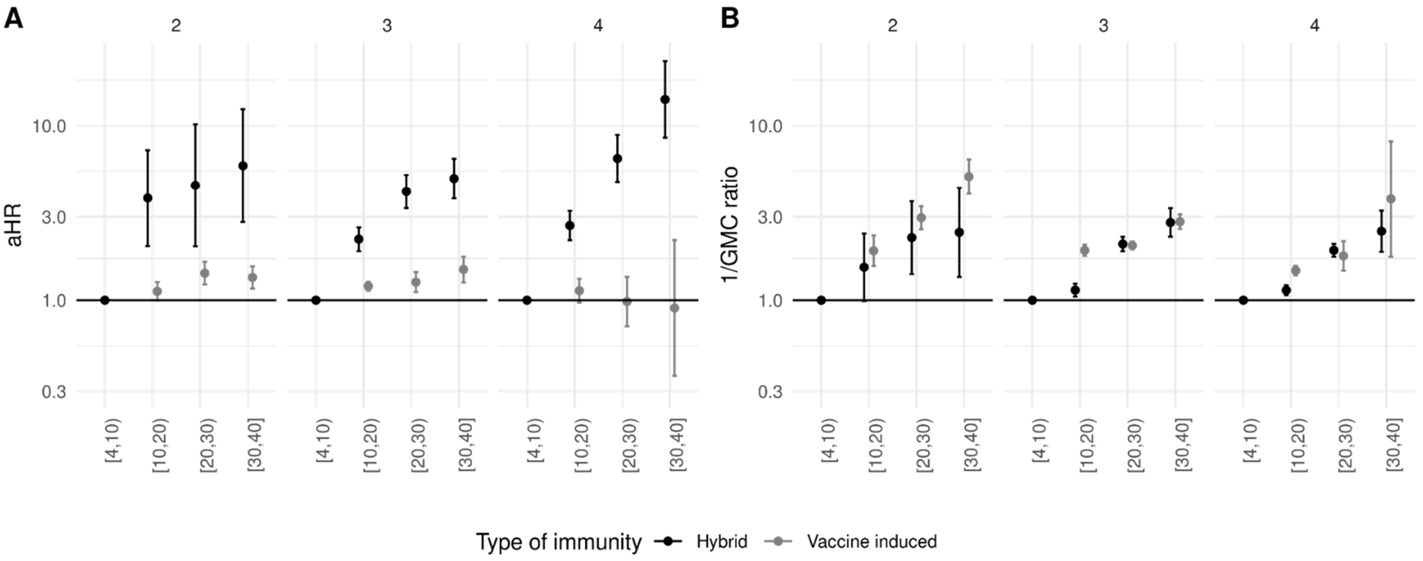


*Figure S2. A. Adjusted hazard ratio (aHR) of infection with Omicron SARS-CoV-2 by type of immunizing events, stratified by the number of immunizing events. B. One divided by the adjusted geometric mean concentration (GMC) ratio of S-antibodies. In both analyses, 4 to 10 weeks after the last immunizing event was the reference group. The group with 4 immunizing events only includes participants aged 60 and older, because younger people were not eligible for 4 vaccinations. Error bars depict 95% confidence intervals.*

*Table S3. Association between anti-S-antibody concentration in quartiles and risk of infection.*

| Quartile log-anti-S-antibody concentration | Person-days | Infections | HR (95% CI) |
|---|---|---|---|
| **(3,8.82]** | 102038 | 321 | [ref] |
| **(8.82,9.43]** | 104379 | 251 | 0.81 (0.69-0.96) |
| **(9.43,10.4]** | 101820 | 203 | 0.65 (0.54-0.78) |
| **(10.4,12.3]** | 103108 | 78 | 0.29 (0.22-0.37) |

*Table S4. Number of person-days and SARS-CoV-2 infections during the study period, hazard ratios (HR) and 95% confidence intervals (CI) for SARS-CoV-2 infection, by number of immunizing events and type of first and last event, among participants with hybrid immunity. Four to 10 weeks after infection as first and vaccination as last event served as the reference group.*

| Number of immunizing events | First and last event, weeks since last event | Person-days | Infections | aHR (95% CI) |
|---|---|---|---|---|
| **3** | Infection Vaccination [4,10) | 49992 | 153 | [ref] |
| **3** | Infection Vaccination [10,20) | 69777 | 227 | 1.3 (1.04-1.63) |
| **3** | Infection Vaccination [20,30) | 52465 | 126 | 1.91 (1.37-2.66) |
| **3** | Infection Vaccination [30,40] | 27461 | 59 | 1.75 (1.22-2.51) |
| **3** | Vaccination Infection [4,10) | 25713 | 20 | 0.5 (0.3-0.81) |
| **3** | Vaccination Infection [10,20) | 48973 | 159 | 1.1 (0.88-1.38) |
| **3** | Vaccination Infection [20,30) | 20187 | 21 | 0.97 (0.6-1.58) |
| **3** | Vaccination Infection [30,40] | 14374 | 15 | 0.79 (0.44-1.42) |
| **3** | Vaccination Vaccination [4,10) | 11397 | 37 | 1.02 (0.71-1.47) |
| **3** | Vaccination Vaccination [10,20) | 15533 | 33 | 1.09 (0.73-1.61) |
| **3** | Vaccination Vaccination [20,30) | 10313 | 15 | 1.01 (0.57-1.8) |
| **3** | Vaccination Vaccination [30,40] | 3777 | 9 | 1.87 (0.93-3.76) |
| **4** | Infection Vaccination [4,10) | 110014 | 272 | [ref] |
| **4** | Infection Vaccination [10,20) | 144460 | 429 | 1.39 (1.16-1.67) |
| **4** | Infection Vaccination [20,30) | 95991 | 169 | 1.86 (1.37-2.53) |
| **4** | Infection Vaccination [30,40] | 42963 | 66 | 2.36 (1.57-3.55) |
| **4** | Vaccination Infection [4,10) | 2930 | 6 | 1.13 (0.5-2.56) |
| **4** | Vaccination Infection [10,20) | 4352 | 20 | 1.59 (1-2.51) |
| **4** | Vaccination Infection [20,30) | 2884 | 6 | 2.61 (1.13-6.04) |
| **4** | Vaccination Infection [30,40] | 1776 | 1 | 0.71 (0.1-5.15) |
| **4** | Vaccination Vaccination [4,10) | 81637 | 142 | 0.81 (0.66-1) |
| **4** | Vaccination Vaccination [10,20) | 102733 | 187 | 1.14 (0.91-1.41) |
| **4** | Vaccination Vaccination [20,30) | 59014 | 70 | 1.29 (0.9-1.85) |
| **4** | Vaccination Vaccination [30,40] | 19684 | 25 | 2.21 (1.32-3.71) |

*Table S5. Number of person-days and SARS-CoV-2 infections during the study period, hazard ratios (HR) and 95% confidence intervals (CI) for SARS-CoV-2 infection, by type of immunizing events, age group, number of immunizing events and weeks since last immunizing event. Four to 10 weeks after the last of three events served as the reference group.*

| Type of immunity, age | Number of immunizing events (weeks after last event) | Person-days | Infections | HR (95% CI) |
|---|---|---|---|---|
| **Hybrid, age 18+** | 2 [4,10) | 12832 | 12 | 0.65 (0.37-1.16) |
| **Hybrid, age 18+** | 2 [10,20) | 20102 | 54 | 2.45 (1.83-3.29) |
| **Hybrid, age 18+** | 2 [20,30) | 11964 | 15 | 2.95 (1.75-4.97) |
| **Hybrid, age 18+** | 2 [30,40] | 6804 | 19 | 4.15 (2.6-6.63) |
| **Hybrid, age 18+** | 3 [4,10) | 210792 | 255 | [ref] |
| **Hybrid, age 18+** | 3 [10,20) | 296881 | 517 | 2.32 (1.99-2.7) |
| **Hybrid, age 18+** | 3 [20,30) | 170143 | 217 | 4.98 (4.11-6.05) |
| **Hybrid, age 18+** | 3 [30,40] | 53892 | 89 | 5.61 (4.39-7.18) |
| **Hybrid, age 18+** | 4 [4,10) | 775376 | 597 | 0.99 (0.85-1.15) |
| **Hybrid, age 18+** | 4 [10,20) | 952282 | 919 | 2.47 (2.12-2.86) |
| **Hybrid, age 18+** | 4 [20,30) | 457110 | 434 | 5.69 (4.74-6.84) |
| **Hybrid, age 18+** | 4 [30,40] | 82836 | 103 | 12.54 (9.65-16.29) |
| **Hybrid, age 18+** | 5 [4,10) | 292148 | 76 | 1.01 (0.78-1.31) |
| **Hybrid, age 18+** | 5 [10,20) | 239877 | 124 | 2.74 (2.18-3.45) |
| **Hybrid, age 18+** | 5 [20,30) | 58151 | 37 | 4.81 (3.35-6.9) |
| **Hybrid, age 18+** | 5 [30,40] | 4337 | 2 | 4.43 (1.1-17.91) |
| **Vaccine-induced, age 18-59** | 2 [4,10) | 103676 | 690 | 1.14 (1.04-1.25) |
| **Vaccine-induced, age 18-59** | 2 [10,20) | 120504 | 781 | 1.32 (1.21-1.45) |
| **Vaccine-induced, age 18-59** | 2 [20,30) | 97541 | 469 | 1.6 (1.42-1.81) |
| **Vaccine-induced, age 18-59** | 2 [30,40] | 52650 | 271 | 1.53 (1.33-1.75) |
| **Vaccine-induced, age 18-59** | 3 [4,10) | 342950 | 2103 | [ref] |
| **Vaccine-induced, age 18-59** | 3 [10,20) | 414089 | 2202 | 1.21 (1.12-1.3) |
| **Vaccine-induced, age 18-59** | 3 [20,30) | 294168 | 1144 | 1.4 (1.24-1.59) |
| **Vaccine-induced, age 18-59** | 3 [30,40] | 123815 | 305 | 1.5 (1.25-1.79) |
| **Vaccine-induced, age 60+** | 2 [4,10) | 4878 | 18 | 1.67 (1.05-2.66) |
| **Vaccine-induced, age 60+** | 2 [10,20) | 7386 | 29 | 1.07 (0.74-1.55) |
| **Vaccine-induced, age 60+** | 2 [20,30) | 10100 | 28 | 1.41 (0.97-2.06) |
| **Vaccine-induced, age 60+** | 2 [30,40] | 23770 | 100 | 1.37 (1.12-1.68) |
| **Vaccine-induced, age 60+** | 3 [4,10) | 678872 | 1635 | [ref] |
| **Vaccine-induced, age 60+** | 3 [10,20) | 762996 | 3696 | 1.15 (1.05-1.26) |
| **Vaccine-induced, age 60+** | 3 [20,30) | 416845 | 889 | 1.11 (0.94-1.31) |

| | | | | |
|---|---|---|---|---|
| **Vaccine-induced, age 60+** | 3 [30,40] | 197409 | 320 | 1.22 (0.99-1.5) |
| **Vaccine-induced, age 60+** | 4 [4,10) | 258106 | 384 | 1.01 (0.85-1.2) |
| **Vaccine-induced, age 60+** | 4 [10,20) | 323388 | 778 | 1.1 (0.93-1.31) |
| **Vaccine-induced, age 60+** | 4 [20,30) | 53415 | 53 | 1.08 (0.78-1.49) |
| **Vaccine-induced, age 60+** | 4 [30,40] | 4090 | 5 | 0.93 (0.38-2.28) |

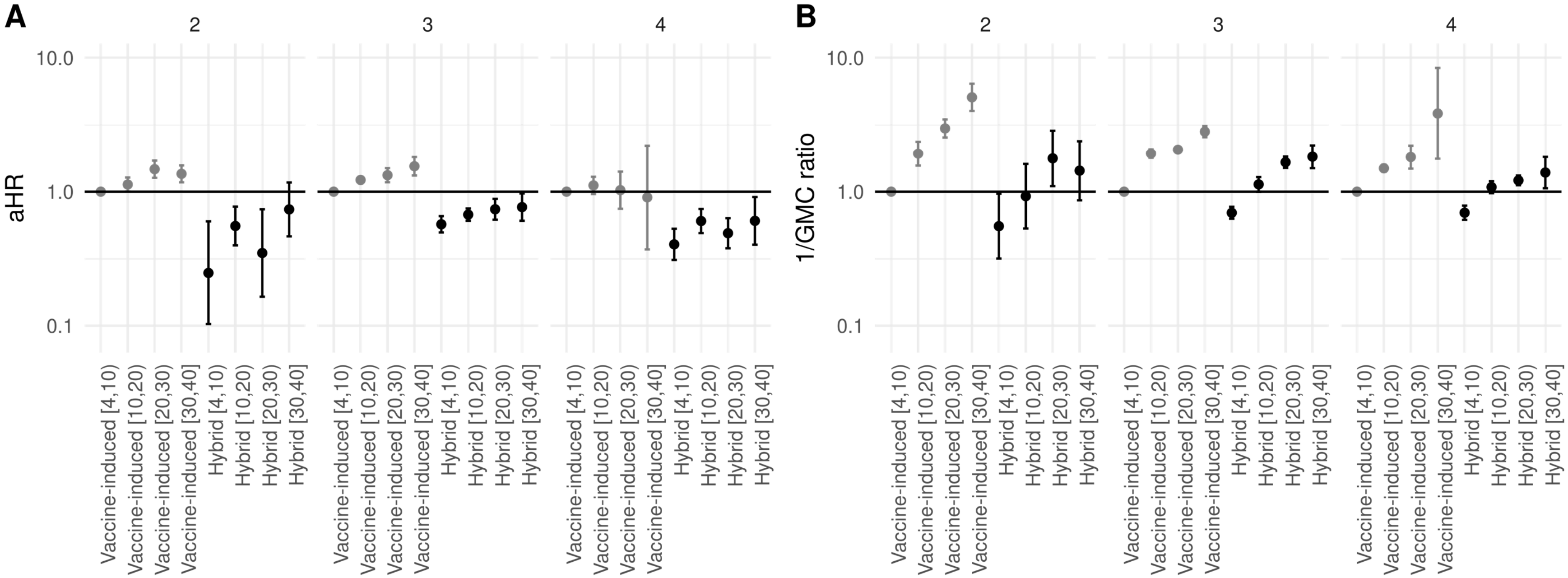


*Figure S3: A. Adjusted hazard ratio (HR) of infection with Omicron SARS-CoV-2 by type of immunizing event, stratified by the number of immunizing events, excluding participants with a previous Omicron infection. B. One divided by the adjusted geometric mean concentration (GMC) ratio of S-antibodies, excluding participants with a previous Omicron infection. In both analyses, 4 to 10 weeks after the last vaccination for vaccine-only immunity was the reference group. The group with 4 immunizing events only includes participants aged 60 and older, because younger people were not eligible for 4 vaccinations. Error bars depict 95% confidence intervals.*
